# Predicting critical illness on initial diagnosis of COVID-19 based on easily-obtained clinical variables: Development and validation of the PRIORITY model

**DOI:** 10.1101/2020.11.27.20237966

**Authors:** Miguel Martinez-Lacalzada, Luis Adrián Viteri-Noël, Luis Manzano, Martin Fabregate-Fuente, Manuel Rubio-Rivas, Sara Luis Garcia, Francisco Arnalich Fernández, José Luis Beato Pérez, Juan Antonio Vargas Núñez, Elpidio Calvo Manuel, Alexia Constanza Espiño Álvarez, Santiago J. Freire Castro, Jose Loureiro-Amigo, Paula Maria Pesqueira Fontan, Adela Pina, Ana María Álvarez Suárez, Andrea Silva Asiain, Beatriz García López, Jairo Luque del Pino, Jaime Sanz Cánovas, Paloma Chazarra Pérez, Gema María García García, Jesús Millán Núñez-Cortés, José Manuel Casas Rojo, Ricardo Gómez Huelgas, for the SEMI-COVID-19 Network

## Abstract

**Objectives:** Currently available COVID-19 prognostic models have focused on laboratory and radiological data obtained following admission. However, these tests are not available on initial assessment or in resource-limited settings. We aim to develop and validate a prediction model, based on clinical history and examination findings on initial diagnosis of COVID-19, to identify patients at risk of critical outcomes.

**Methods:** We used data from the SEMI-COVID-19 Registry, a nationwide multicenter cohort of consecutive patients hospitalized for COVID-19 from 132 centers in Spain. Clinical signs and symptoms, demographic variables, and medical history ascertained at hospital admission were screened using Least Absolute Shrinkage and Selection Operator (LASSO) and logistic regression to construct a predictive model. We externally validated the final model in a separate cohort of patients admitted at less-complex hospitals (< 300 beds).We undertook decision curve analysis to assess the clinical usefulness of the predictive model. The primary outcome was a composite of in-hospital death, mechanical ventilation or admission to intensive care unit.

**Results:** There were 10,433 patients, 7,850 (primary outcome 25.1%) in the development cohort and 2,583 (primary outcome 27.0%) in the validation cohort. Variables in the final model included: age, cardiovascular disease, chronic kidney disease, dyspnea, tachypnea, confusion, systolic blood pressure, and SpO_2_≤93% or oxygen requirement.The C-statistic in the development cohort was 0.823 (95% CI,0.813-0.834). On external validation, the C-statistic was 0.792 (95% CI,0.772-0.812). The model showed a positive net benefit for threshold probabilities between 3% and 79%.

**Conclusions:** Among patients presenting with COVID-19, the model based on easily-obtained clinical information had good discrimination and generalizability for identifying patients at risk of critical outcomes without the need of additional testing. The online calculator provided would facilitate triage of patients during the pandemic. This study could provide a useful tool for decision-making in health systems under pandemic pressure and resource-limited settings.

## INTRODUCTION

The clinical spectrum of severe acute respiratory syndrome coronavirus 2 (SARS-CoV-2) infection ranges from an asymptomatic state to critical illness; the symptomatic profile is called coronavirus disease 2019 or COVID-19 [1, 2]. As of January 12, 2021, the COVID-19 pandemic has affected more than 88 million people worldwide, and has led to over 1.9 million deaths [3]. Notably, Spain has been one of the countries with the highest number of patients with COVID-19 [4]. To optimize the use of limited healthcare resources, it would be essential to identify, as early as possible, those patients who are at high risk of progressing to critical illness that necessitates admission to intensive care unit (ICU) or mechanical ventilation, or that may lead to mortality.

To date, studies of COVID-19 prognostic factors have focused on laboratory measurements and radiological examinations obtained following admission [5-15], which are not available in outpatient or resource-limited settings. Recently published well-developed models tend not to include clinical variables obtained from history and examination carried out on initial assessment [9-13]. Where one machine learning model has addressed basic clinical features, it has narrowed down the prediction to the mortality outcome only and lacks wider generalizability [16]. Furthermore, a critical appraisal of the COVID-19 models has shown poor reporting and high risk of bias [14].

Prediction models based on easy-to-collect data without using imaging or laboratory measures have previously been developed for other infectious diseases, e.g. meningitis and pneumonia [17-19]. As a global health emergency, management of COVID-19 would benefit from a prediction model that can be readily applied on initial diagnosis. Therefore, we developed and externally validated a prediction model, based on easily obtained clinical measures at presentation with confirmed COVID-19 diagnosis, to identify patients at risk of developing critical outcomes.

## METHODS

### Study design and data source

This study was based on the SEMI (Sociedad Española de Medicina Interna) COVID-19 Registry [20]. It is an ongoing multicenter nationwide cohort of consecutive patients hospitalized for COVID-19 across Spain. Patients were confirmed to be COVID-19 cases defined as a positive result on real-time reverse-transcription-polymerase-chain-reaction (RT-PCR) for the presence of SARS-CoV-2 in nasopharyngeal swab specimens or sputum samples. Exclusion criteria were age under 18 years, subsequent admissions and refusal or withdrawal of informed consent. Clinical baseline data, history of previous medication, known comorbidities, laboratory and imaging variables were collected on admission. In addition, treatments and complications during hospitalization, status on day of discharge and/or 30 days after diagnosis were obtained. Registry’s characteristics have been previously described in detail [20]. The SEMI-COVID-19 Registry was approved by the Provincial Research Ethics Committee of Málaga (Spain) and by Institutional Research Ethics Committees of each participating hospital.

For the study, we used data from patients admitted in 132 hospitals between March 23 and May 21, 2020. Development and validation cohorts were defined according to the size of hospitals. Model development was performed on a cohort of patients from hospitals with at least 300 beds, and validated on a separate cohort from hospitals with less than 300 beds. This approach was taken to examine the external validity of the prognostic model [21] in a lower complexity level setting compared to the development setting [22]. The study was reported following the TRIPOD (Transparent Reporting of a multivariable prediction model for Individual Prognosis or Diagnosis) guidelines (Supplementary Table S1) [23].

### Outcome description

Primary outcome, critical illness during hospitalization, was defined as the composite of in-hospital death, mechanical ventilation or admission to intensive care unit (ICU), according to previously published studies [10, 24, 25].

### Potential predictors

To develop a predictive model based only on easily measurable variables registered at admission, from the available variables at SEMI-COVID-19 Registry we only considered clinical signs and symptoms, demographic variables, and medical history. Our approach for selecting predictors was developed to meet the recommendation that new prediction models, rather than using purely data-driven selection, should build on previous literature and expert opinion [14].

An initial list of 29 candidate variables was selected based on review of the existing evidence [5-16], clinical plausibility and relevance to clinical care. Demographic variables included age, sex, ethnicity (defined as Caucasian, Latino or others), history of smoking and previous medication as angiotensin converting enzyme inhibitors (ACEi) and angiotensin receptor blockers (ARBs). Medical history included hypertension, cardiovascular disease (history of cerebrovascular disease, peripheral arterial disease, myocardial infarction, angina pectoris, heart failure or atrial fibrillation), moderate or severe dependency for activities of daily living (Barthel index score <60), diabetes mellitus, obesity and chronic respiratory diseases (asthma, chronic obstructive pulmonary disease, obstructive sleep apnea or hypopnea syndrome), severe chronic kidney disease (previous serum creatinine level >3 mg/dl or history of dialysis), malignancy (solid tumor, leukemia or lymphoma), chronic liver disease, immunocompromised status (autoimmune diseases, solid-organ transplant recipients, HIV infection or previous immunosuppressive treatment including systemic steroids). Clinical signs and symptoms were cough, arthromyalgia, ageusia/anosmia, asthenia/anorexia, headache, gastrointestinal symptoms, fever (temperature ≥38°C or history of fever), systolic blood pressure, heart rate, tachypnea (respiratory rate >20 breaths/min), pulmonary rales, confusion, dyspnea, and peripheral oxygen saturation (SpO_2_) ≤93% at room air or supplementary oxygen requirement at admission [26].

To improve consensus on model applicability, a 1-round online questionnaire was conducted among a multidisciplinary panel of 24 physicians involved in COVID-19 clinical management at nursing homes, emergency departments, primary care centers and hospitalization wards (6 per each setting). The panelists were asked to rate (on a 9-point Likert scale) the availability/reliability of each predictor, its ability to predict the outcome, the best way to merge predictors of rare occurrence, and the maximum number of variables this model should contain. Agreement was considered when ≤7 panelists rated outside the 3-point region containing the median [27].

### Statistical analysis

The predictive model, called PRIORITY, was presented as the formula for estimating the probability of COVID-19 critical illness outcome, as well as an associated web-based calculator. Patients’ characteristics were summarized in terms of frequencies and percentages and by the mean and standard deviation (SD). Statistical analysis was performed using R v4.0.0, with the mice, mfp, glmnet, pROC, and rmda packages.

#### Model development

Missing values in the potential predictors were imputed using single imputation, a reasonable alternative to multiple imputation when dealing with relatively few missings [28]. A stochastic single imputation dataset was created for both cohorts (development and validation) as the first of a series of datasets generated through multiple imputation by chained equations. Quantitative variables were kept as continuous to avoid loss of prognostic information, and non-linear relationships were modelled by multivariate fractional polynomials with a maximum of 2 degrees of freedom [29]. The least absolute shrinkage and selection operator (LASSO) method (30) was used to identify a parsimonious set of potential predictors of critical illness. We selected the regulation penalty parameter that minimized the 10-fold cross-validation mean squared error for a maximum number of predictive features in the model settled by the expert panel agreement. Then, this subset of predictors was entered into a logistic regression model, and those statistically significant (p<0.05) were retained. The model coefficients were represented as odds ratios (OR), and 95% confidence intervals (95% CI) were obtained using 1000 bootstrap samples.

#### Model performance

We used Nagelkerke’s R^2^ to evaluate the overall predictive accuracy of the model. The overall discriminatory ability was assessed using the C-statistic, as the area under the receiver operating characteristic curve (AUC ROC), with 95% CI by stratified bootstrap resampling. Calibration of the model was assessed graphically, and an overfitting-corrected estimate of the calibration slope was derived by bootstrapping 1000 resamples [31].

#### Model validation

Internal validation of the model was assessed by 10-fold cross-validation [30]. We externally validated the final model in a separate cohort of patients admitted at less-complex hospitals (< 300 beds) to assess model generalizability [21].

#### Sensitivity analysis

To assess the impact of assumptions adopted in the model development, we carried out a complete-case analysis, using only those patients with complete data in the potential predictors. We also developed models without restricting the maximum number of predictors or using linear continuous predictors instead of fractional polynomial terms.

#### Decision curve analysis

We undertook decision curve analysis (DCA) to assess the clinical usefulness of the predictive model when used to prioritize hospital referrals that are most likely to require critical care [32]. For the whole range of decision threshold probabilities (*p*_*t*_), the net benefit (NB) of the model was compared to default strategies of treat (or refer) all or no patients. The NB was calculated as: True positives/N–(False positives/N)*(*p*_*t*_ /(1-*p*_*t*_), with N total sample size, and represented in a decision curve plot. The benefit was also quantified in terms of reduction in avoidable hospitalization referrals per 100 patients as: (NB of the model – NB of treat all)/(p_t_ /(1-p_t_))×100 [32].

The choice of *p*_*t*_ will vary across different regions, according to changing epidemiological situations and availability of health resources. At a low threshold, false negatives are minimized at the expense of unnecessary referrals. At a high threshold, patients would be referred less frequently, but some high-risk patients may not be derived to the hospital.

## RESULTS

From a total of 11,523 patients of the SEMI-COVID-19 Registry, 10,433 were considered in this study. The development cohort included 7,850 (75.2%) patients, of which 1,967 (25.1%) presented critical illness (650 [8.3%] admitted to the ICU and 1,598 [20.4%] died). The mean age was 65.8 ± 16.4 years (57.2% male), and 66.5% presented comorbidities. Demographics and clinical characteristics for the development cohort are shown on *Table 1*.

**Table 1.**
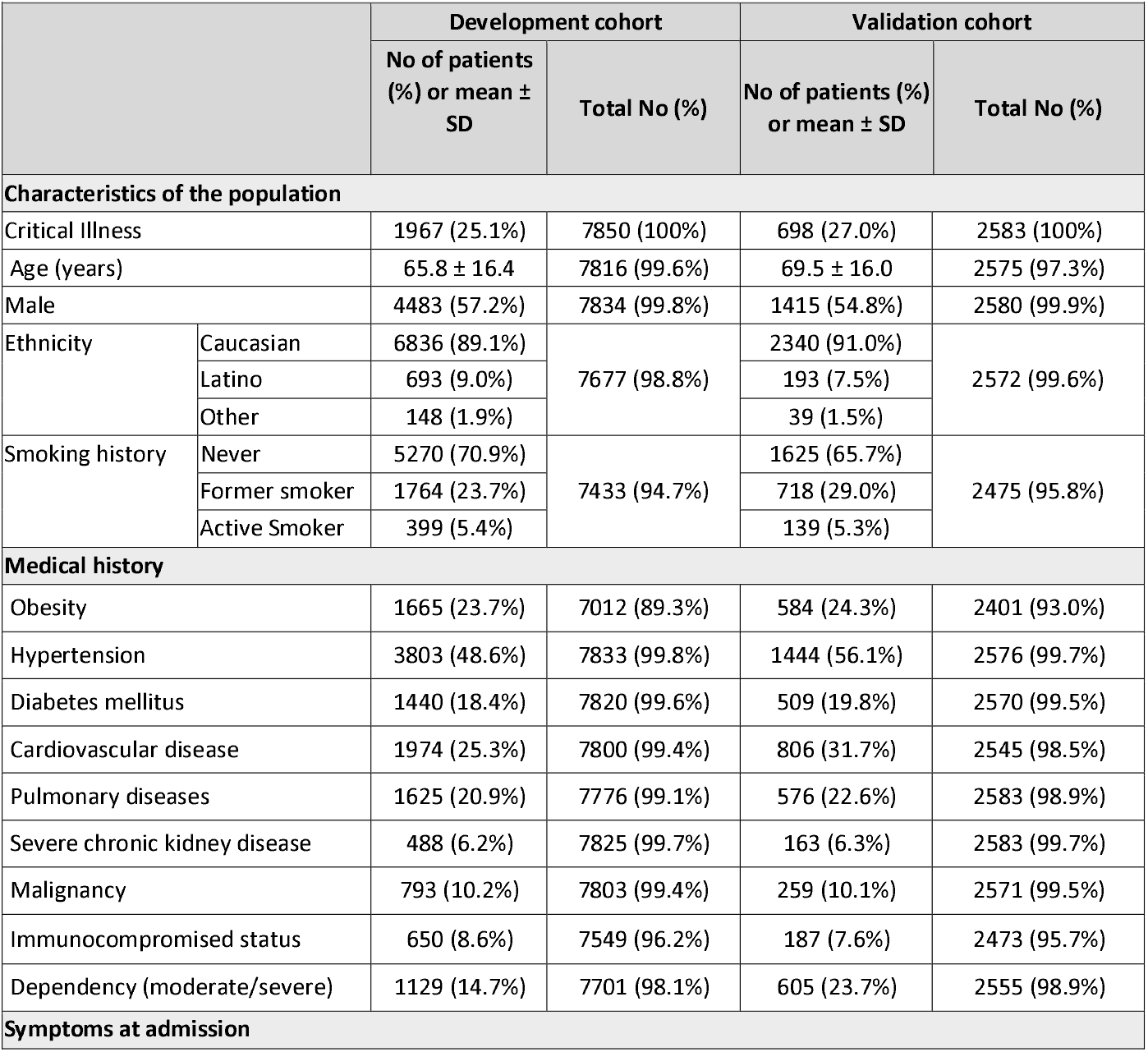
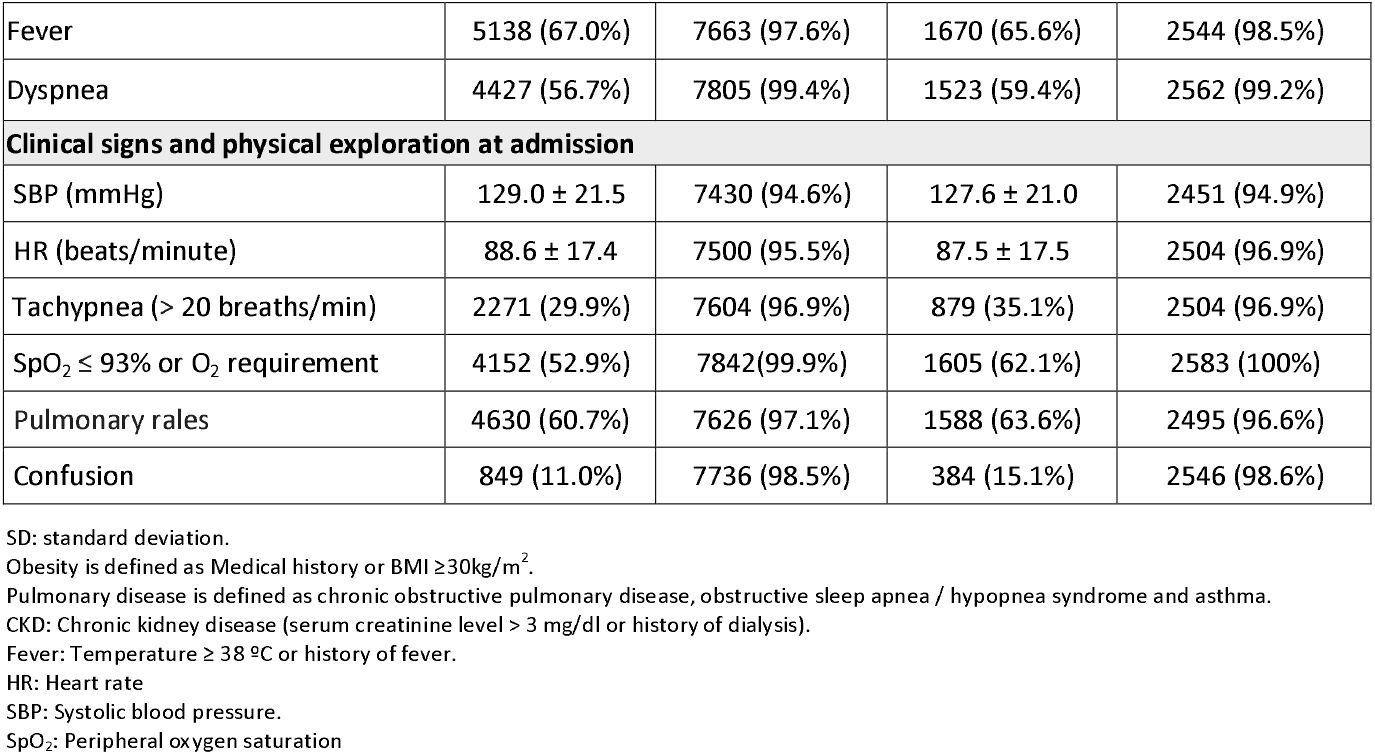
**Demographics and clinical characteristics among patients included in the development cohort and validation cohort.**

|  |  | Development cohort |  | Validation cohort |  |
| --- | --- | --- | --- | --- | --- |
|  |  | No of patients (%) or mean ± SD | Total No (%) | No of patients (%) or mean ± SD | Total No (%) |
| Characteristics of the population |  |  |  |  |  |
| Critical Illness |  | 1967 (25.1%) | 7850 (100%) | 698 (27.0%) | 2583 (100%) |
| Age (years) |  | 65.8 ± 16.4 | 7816 (99.6%) | 69.5 ± 16.0 | 2575 (97.3%) |
| Male |  | 4483 (57.2%) | 7834 (99.8%) | 1415 (54.8%) | 2580 (99.9%) |
| Ethnicity | Caucasian | 6836 (89.1%) | 7677 (98.8%) | 2340 (91.0%) | 2572 (99.6%) |
|  | Latino | 693 (9.0%) |  | 193 (7.5%) |  |
|  | Other | 148 (1.9%) |  | 39 (1.5%) |  |
| Smoking history | Never | 5270 (70.9%) | 7433 (94.7%) | 1625 (65.7%) | 2475 (95.8%) |
|  | Former smoker | 1764 (23.7%) |  | 718 (29.0%) |  |
|  | Active Smoker | 399 (5.4%) |  | 139 (5.3%) |  |
| Medical history |  |  |  |  |  |
| Obesity |  | 1665 (23.7%) | 7012 (89.3%) | 584 (24.3%) | 2401 (93.0%) |
| Hypertension |  | 3803 (48.6%) | 7833 (99.8%) | 1444 (56.1%) | 2576 (99.7%) |
| Diabetes mellitus |  | 1440 (18.4%) | 7820 (99.6%) | 509 (19.8%) | 2570 (99.5%) |
| Cardiovascular disease |  | 1974 (25.3%) | 7800 (99.4%) | 806 (31.7%) | 2545 (98.5%) |
| Pulmonary diseases |  | 1625 (20.9%) | 7776 (99.1%) | 576 (22.6%) | 2583 (98.9%) |
| Severe chronic kidney disease |  | 488 (6.2%) | 7825 (99.7%) | 163 (6.3%) | 2583 (99.7%) |
| Malignancy |  | 793 (10.2%) | 7803 (99.4%) | 259 (10.1%) | 2571 (99.5%) |
| Immunocompromised status |  | 650 (8.6%) | 7549 (96.2%) | 187 (7.6%) | 2473 (95.7%) |
| Dependency (moderate/severe) |  | 1129 (14.7%) | 7701 (98.1%) | 605 (23.7%) | 2555 (98.9%) |
| Symptoms at admission |  |  |  |  |  |

|  |  |  |  |  |
| --- | --- | --- | --- | --- |
| Fever | 5138 (67.0%) | 7663 (97.6%) | 1670 (65.6%) | 2544 (98.5%) |
| Dyspnea | 4427 (56.7%) | 7805 (99.4%) | 1523 (59.4%) | 2562 (99.2%) |
| **Clinical signs and physical exploration at admission** |  |  |  |  |
| SBP (mmHg) | 129.0 ± 21.5 | 7430 (94.6%) | 127.6 ± 21.0 | 2451 (94.9%) |
| HR (beats/minute) | 88.6 ± 17.4 | 7500 (95.5%) | 87.5 ± 17.5 | 2504 (96.9%) |
| Tachypnea (> 20 breaths/min) | 2271 (29.9%) | 7604 (96.9%) | 879 (35.1%) | 2504 (96.9%) |
| SpO<sub>2</sub> ≤ 93% or O<sub>2</sub> requirement | 4152 (52.9%) | 7842 (99.9%) | 1605 (62.1%) | 2583 (100%) |
| Pulmonary rales | 4630 (60.7%) | 7626 (97.1%) | 1588 (63.6%) | 2495 (96.6%) |
| Confusion | 849 (11.0%) | 7736 (98.5%) | 384 (15.1%) | 2546 (98.6%) |

SD: standard deviation.
Obesity is defined as Medical history or BMI ≥30kg/m<sup>2</sup>.
Pulmonary disease is defined as chronic obstructive pulmonary disease, obstructive sleep apnea / hypopnea syndrome and asthma.
CKD: Chronic kidney disease (serum creatinine level > 3 mg/dl or history of dialysis).
Fever: Temperature ≥ 38 °C or history of fever.
HR: Heart rate
SBP: Systolic blood pressure.
SpO<sub>2</sub>: Peripheral oxygen saturation

### Model development and performance

From an initial list of 29 candidate variables, the expert panel forged an agreement on 21 potential predictors for further evaluation in the predictive model. So, chronic liver disease, previous medication (ACEi and ARBs), cough, arthromyalgia, ageusia/anosmia, asthenia/anorexia, headache, gastrointestinal symptoms were excluded. Moreover, consensus was achieved for including a range between 5 and 9 variables in the final model. These 21 potential predictors were included in the LASSO predictor selection process. A subset of 9 variables were retained as the best predictors of critical illness (*Supplementary Figure S1*), including age squared, moderate or severe dependency, cardiovascular disease, moderate or severe chronic kidney disease, dyspnea, tachypnea, confusion, reciprocal of systolic blood pressure squared, and SpO_2_ ≤ 93% or oxygen requirement. A multivariable logistic regression model was then fitted with these 9 variables. All of them, except for moderate or severe dependency, were statistically significant (*Table 2*).

**Table 2.**
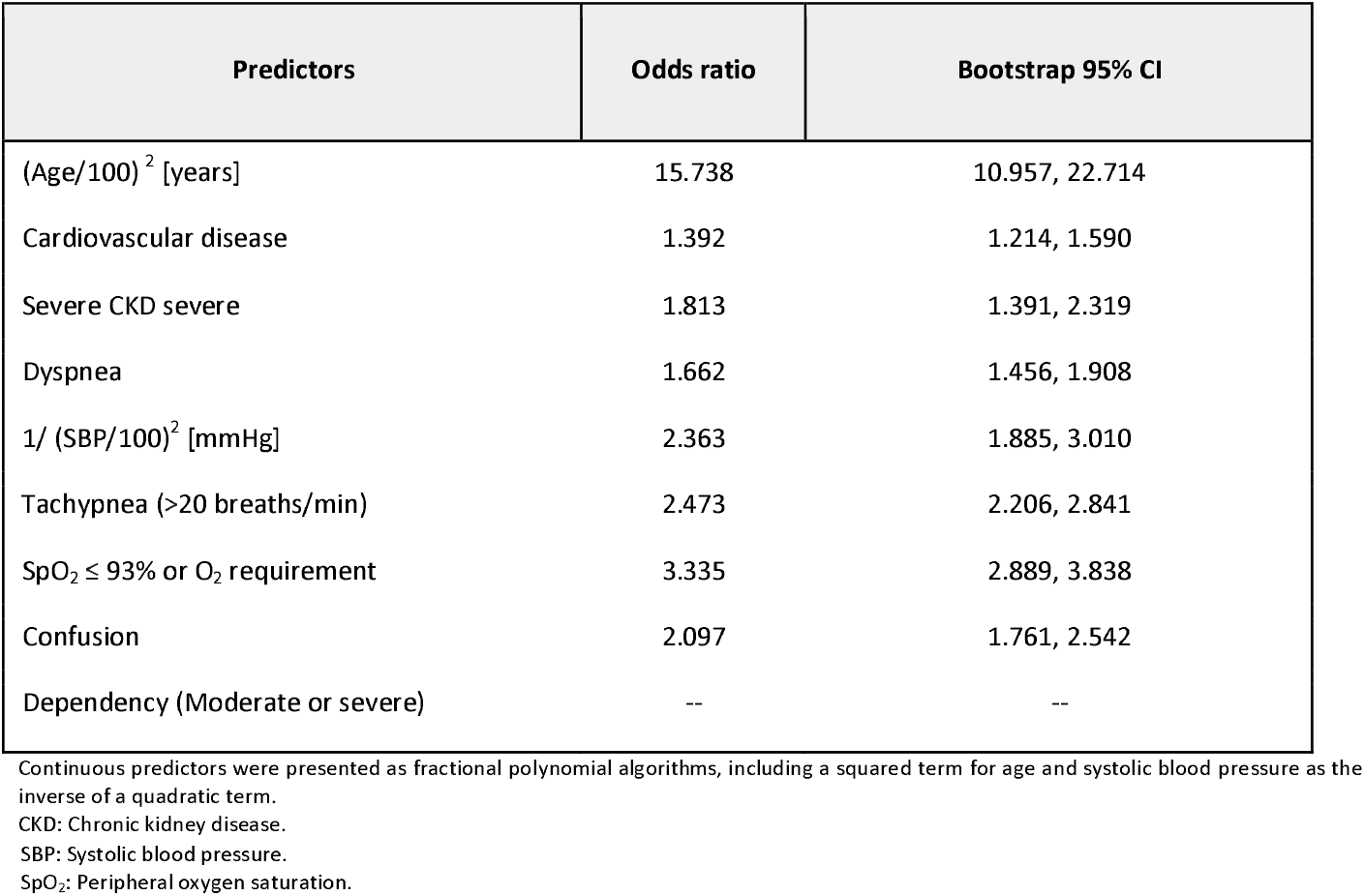
Multivariable LASSO logistic regression of critical illness prediction in COVID-19.

|  |  |  |
| --- | --- | --- |
| **Predictors** | **Odds ratio** | **Bootstrap 95% CI** |
| (Age/100)<sup>2</sup> [years] | 15.738 | 10.957, 22.714 |
| Cardiovascular disease | 1.392 | 1.214, 1.590 |
| Severe CKD severe | 1.813 | 1.391, 2.319 |
| Dyspnea | 1.662 | 1.456, 1.908 |
| 1/ (SBP/100)<sup>2</sup> [mmHg] | 2.363 | 1.885, 3.010 |
| Tachypnea (>20 breaths/min) | 2.473 | 2.206, 2.841 |
| SpO<sub>2</sub> ≤ 93% or O<sub>2</sub> requirement | 3.335 | 2.889, 3.838 |
| Confusion | 2.097 | 1.761, 2.542 |
| Dependency (Moderate or severe) | -- | -- |

Continuous predictors were presented as fractional polynomial algorithms, including a squared term for age and systolic blood pressure as the inverse of a quadratic term.
CKD: Chronic kidney disease.
SBP: Systolic blood pressure. SpO<sub>2</sub>: Peripheral oxygen saturation.

Based on the logistic regression model, the probability of critical COVID-19 illness could be calculated as:

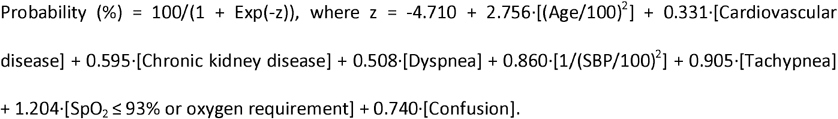

All predictors were coded as binary variables (1 when present and 0 when absent) except for age (years) and systolic blood pressure (mmHg). We also developed an online calculator based on this model (*Supplementary Figure S2*), that is accessible at https://www.evidencio.com/models/show/2344.

The final model had an R^2^ of 0.346 and an apparent C-statistic of 0.823 (95% CI 0.812, 0.833) to discriminate between patients with and without critical illness in the development cohort (*Figure 1a*). By internal 10-fold cross-validation, estimated performance of the model was 0.822 (95% CI 0.789, 0.848). After bootstrap resampling, the agreement between the observed outcomes and predicted probabilities in the development cohort showed good calibration with a slope of 0.995 (*Supplementary Figure S3a*).

**Figure 1.**
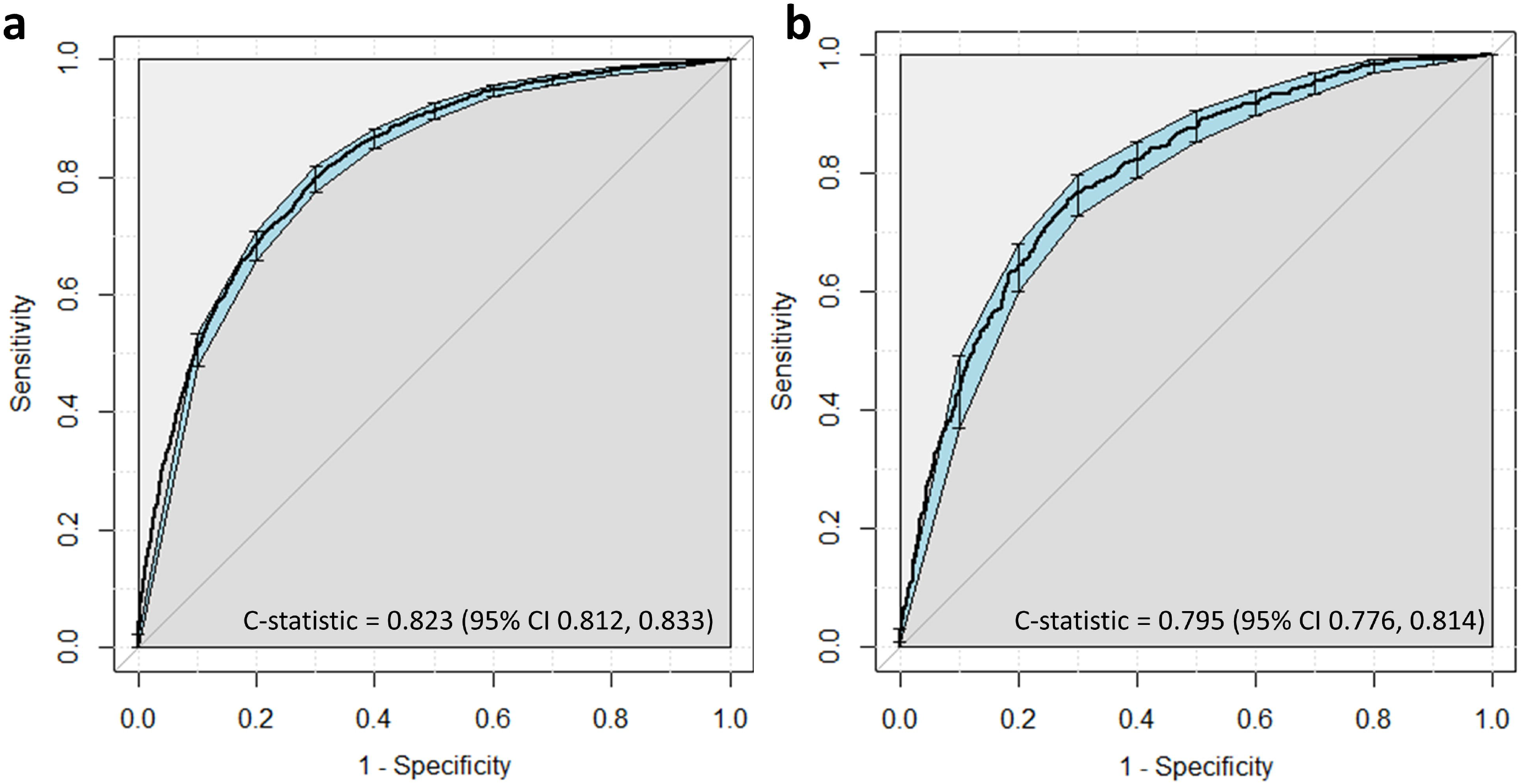
Area under the receiver-operator characteristic curve (AUC ROC) of the predictive model for critical illness among patients hospitalized with COVID-19. (a). AUC ROC in the development cohort, n=7850 patients from hospitals with equal or more than 300 beds. (b). AUC ROC in the validation cohort, n=2583 patients from hospitals with less than 300 beds. 95% coefficient intervals (CI) computed with 1000 bootstrap replicates.

### External validation

The validation cohort included 2583 (24.8%) patients, of which 698 (27.0%) presented critical illness (200 [7.7%] admitted to the ICU and 594 [23.0%] died). The mean age was 69.5 ± 16.0 years, (54.8% male), and 69.8% presented comorbidities (*Table 1*). The final model showed good discrimination when externally validated, with a C-statistic of 0.795 (95% CI 0.776, 0.814) (*Figure 1b*), and a calibration slope of 0.883 (*Supplementary Figure S3b*).

### Sensitivity analysis

We carried out a complete-case analysis selecting as development cohort the 5513 patients with complete data on the 21 potential predictors and the outcome. The resulting model had the same predictors as the final model with imputed data. R^2^ was 0.324, with an apparent C-statistic of 0.813 (95% CI 0.800, 0.823) and a slope of 0.992. Next, we fitted a new model with no restriction in maximum number of predictors, resulting in a model with 15 variables, adding sex, moderate or severe dependency, diabetes mellitus, malignancy, immunocompromised status, pulmonary rales and heart rate cubed, to the 8 predictors in the PRIORITY model. R^2^ was 0.348, with a C-statistic of 0.832 (95% CI 0.821, 0.842) and a calibration slope of 0.995. Likewise, we fitted an alternative model using linear continuous predictors instead of fractional polynomial terms. The linear term of systolic blood pressure was not found to be a significant predictor, while moderate or severe dependency was included in the model. R^2^ was 0.339, C-statistic of 0.819 (95% CI 0.809, 0.830) and a slope of 0.996.

### Net benefit of model use

The decision curve analysis (*Figure 2*) showed a positive NB for *p*_*t*_ between 3% and 79%, compared to default strategies (treat-all or treat-none). For low thresholds, below 3%, the NB of the model was comparable to managing all COVID-19 patients as if they will progress to critical illness (treat-all strategy). *Table 3* presents estimates of the NB of using the model and the reduction in avoidable hospitalization referrals for different *p*_*t*_.

**Table 3.** **Net benefit of using the PRIORITY prediction model compared to managing COVID-19 patients assuming that all of them will progress to critical illness.**

| Probability threshold (p, %) | Net benefit |  | Advantage of using the PRIORITY model |  |
| --- | --- | --- | --- | --- |
|  | Treat all | Prediction Model | Difference in the net benefit | Reduction in avoidable hospitalization referrals per 100 patients |
| 1 | 0.243 | 0.243 | 0 | 0 |
| 2 | 0.235 | 0.235 | 0 | 0 |
| 3 | 0.227 | 0.227 | 0 | 0 |
| 4 | 0.219 | 0.220 | 0.001 | 2 |
| 5 | 0.211 | 0.214 | 0.003 | 6 |
| 10 | 0.167 | 0.188 | 0.021 | 19 |
| 15 | 0.118 | 0.164 | 0.046 | 26 |
| 20 | 0.063 | 0.144 | 0.081 | 32 |
| 25 | 0.001 | 0.122 | 0.121 | 36 |
| 50 | -0.499 | 0.005 | 0.504 | 50 |
| 75 | -1.998 | 0.004 | 2.002 | 67 |
| 79 | -2.569 | 0 | 2.569 | 68 |
| 99 | -73.943 | 0 | 73.943 | 75 |

Net benefit: percentage of true positives minus the percentage of false positives weighted by the ratio ( $pt/(1-pt)$ ). Reduction in avoidable hospitalization referrals per 100 patients: $(\text{net benefit of the model} - \text{net benefit of treat all})/(pt/(1-pt)) \times 100$ .

**Figure 2.**
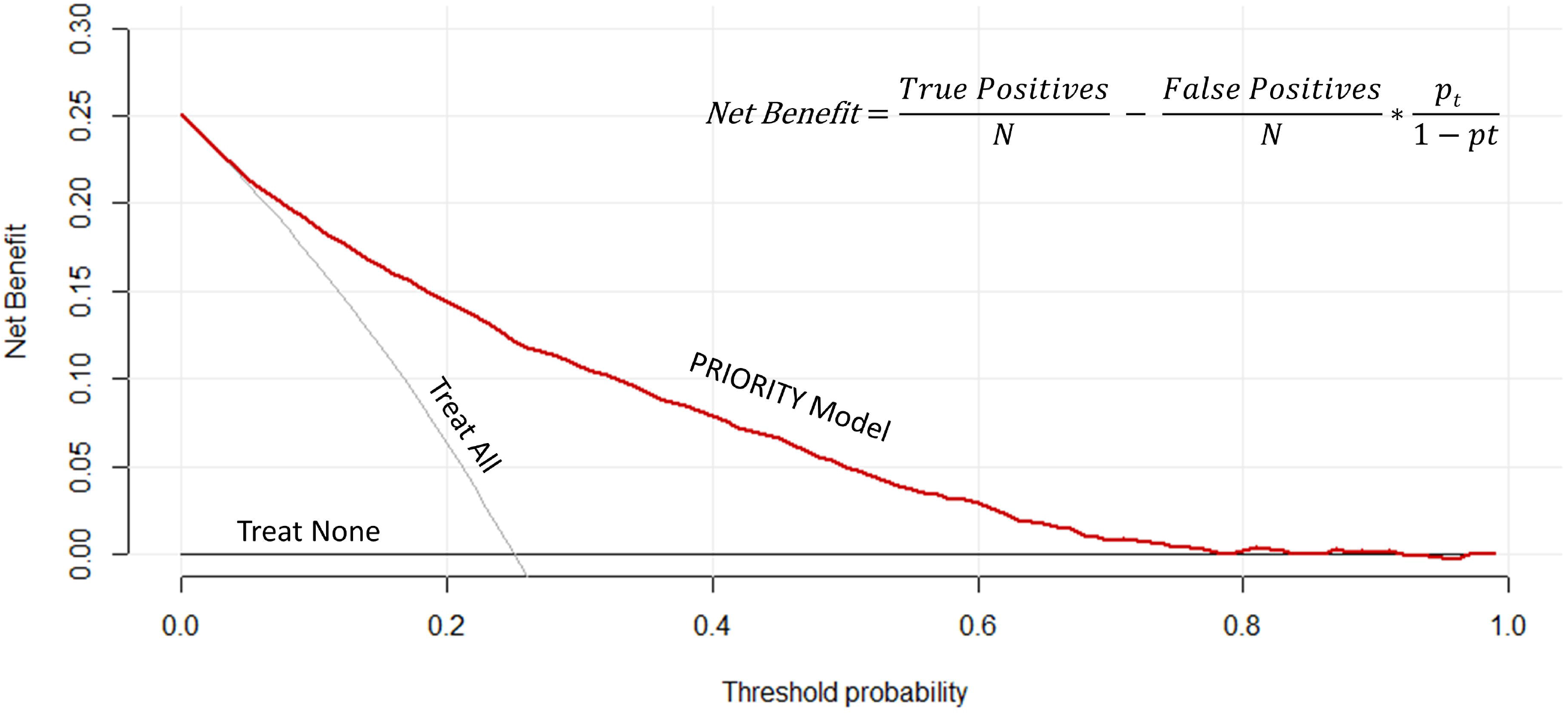
Decision curves of the predictive model for severe COVID-19. The x-axis represents threshold probabilities and the y-axis the net benefit.

## DISCUSSION

We developed and validated a new clinical risk model to predict COVID-19 critical illness based on eight simple clinical features easily available on initial assessment, which would be useful in resource-limited or out-of-hospital settings without access to other complementary tests. The model was well calibrated, had good discrimination, and performed robustly in an external validation cohort. Moreover, it showed a potential clinical benefit in a variety of scenarios covering different healthcare situations over a range of threshold probabilities, highlighting its practical usefulness. Its web-based calculator can facilitate its immediate application for frontline clinicians.

This study has several methodological strengths maximizing internal and external validity [23]. To the best of our knowledge, this is the first generalizable COVID-19 predictive model built with simple clinical information excluding imaging and laboratory data. We developed and validated the model in a large multicenter, national cohort. Ours was a cohort twice as large as the previous model using simple information [16]. Our model excluded readmissions, a feature that focuses the analysis on the question of interest, i.e. the need of triage in patients at their first COVID-19 presentation. The methodology was rigorous, avoiding data-driven predictor selection and biases that affected previous studies [14]. The practical application of the model was maximized by forging an agreement among an expert panel on key issues. The performance of the model was good, which allowed discrimination between progression or not to COVID-19 critical illness. Moreover, the model was validated in a separate cohort of patients admitted in smaller hospitals, showing strong reproducibility in a healthcare setting of a different complexity level [21, 22]. Our decision curve analysis showed that the model could be useful for triage of patients under pandemic pressure, providing underpinning evidence to guide policymakers’ decisions.

The strength of our findings should be interpreted in light of some limitations. Although we carefully selected easily available clinical and demographic variables that could be potentially applied in settings without access to laboratory or radiology tests, the data were collected at the time of hospital admission, which represents an important selection bias that would require further studies in an outpatient setting. We used data collected in a situation of healthcare pressure due to the pandemic peak, so the data quality may be variable across centers. In this regard, it is notable that missing data per predictor variable were relatively low. To reduce the impact of data loss we used imputation. The sensitivity analysis showed that the assumptions adopted in the model development, such as data imputation, restricting the number of predictors or modelling non-linear relationships, did not impact in the model performance. Considering the balance between strengths and limitations, our model is ready for application as a triage tool within the context of an evaluative study to allow solidification of evidence about model effectiveness in practice.

An external validation study [15] of 22 previously published prognostic models showed that oxygen saturation and age were the most discriminating univariate predictors for in-hospital mortality, and that none of the multivariate models had superior performance than these individual predictors. It is important to point out that the PRIORITY model, despite its simplicity, showed a performance similar to previously published models including laboratory and imaging data [9-16]. For example, our model (C-statistic 0.82) would be expected to dominate in health economic terms the model of Knight et al [12] (C-statistic 0.79) on the basis that it would not incur costs involved in imaging and laboratory tests.

Our model could be applied in triage, using easily measurable variables available in settings without access to laboratory or radiology tests, identifying high-risk patients for referral to hospital. The DCA provides information to underpin clinical management and policy-making under COVID-19 pandemic pressure. The PRIORITY model has potential value, resulting in higher net benefit than the default strategies (hospitalize all or hospitalize none), over a range of risk thresholds which could be considered as relevant in clinical practice. For example, in situations under pandemic peak pressure or low-resource healthcare systems, policy-makers may consider a cut-off point up to 20%, a threshold that will be associated with higher reduction in unnecessary critical care admissions. However, in situations with low numbers of COVID-19 cases and little risk of overwhelming the critical care capacity, a lower threshold may be considered. For example, a 5% cut-off could be appropriate to make decisions on early referral to hospital attention. We recommend objectively defining specific cut-off points considering the circumstances and the availability of health resources.

In summary, we developed and validated a new prediction model, called PRIORITY, to estimate the risk of critical illness in patients with COVID-19, based on eight clinical variables easily measurable in resource-limited or out-of-hospital settings without access to other complementary tests. This model could help in triage of patients at risk for critical COVID-19 illness. The study provides underpinning evidence to inform decision-making in health systems under pandemic pressure.

## Data Availability

The data that support the findings of this study are available on request from the SEMI-COVID-19 Registry Coordinating Center, S&H Medical Science Service and the corresponding author, [LM].

## ACKNOWLEDGEMENTS

We gratefully acknowledge all the investigators who participate in the SEMI-COVID-19 Registry, especially those from Hospital Universitario Ramón y Cajal (Luis F. Abrego-Vaca, Ana Andreu-Arnanz, Octavio A. Arce-García, Marta Bajo-González, Pablo Borque-Sanz, Alberto Cózar-Llistó, Beatriz Del Hoyo-Cuenda, Alejandra Gamboa-Osorio, Isabel García-Sánchez, Óscar A. López-Cisneros, Borja Merino-Ortiz, Elisa Riera-González, Jimena Rey-García, Cristina Sánchez-Díaz, Grisell Starita-Fajardo, Cecilia Suárez-Carantoña, Svetlana Zhilina Zhilina). We especially thank to our colleagues at Hospital Universitario Ramon y Cajal, IRYCIS: Nuria Bara Ledesma, Andrés González García and José Luis Calleja López (Internal Medicine Dept.), and Javier Zamora and Borja M. Fernandez-Felix (Clinical Biostatistics Unit), for their valuable contributions to the review of the manuscript. The authors would also like to gratefully acknowledge Professor Khalid S. Khan, Distinguished Investigator at the University of Granada, Spain, for his support and advice on the manuscript. Finally, we also thank the SEMI-COVID-19 Registry Coordinating Center, S&H Medical Science Service, for their quality control data, logistic and administrative support. The authors declare that there are no conflicts of interest.

## AUTHORS CONTRIBUTIONS

MML, LAVN and MFF planned, conceived the study, analysed and interpreted the data. MML, LAVN, MFF and LM wrote the original draft of the manuscript. MRR, SLG, FAF, JLBP, JAVN, ECM, ACEA, SJFC, JLA, PMPF, AP, AMAS, ASA, BGL, JLP, JSC, PCP, GMGG, JMNC, JMCR and RGH contributed to read and approved the final version of the manuscript. MML and LAVN are joint first authors. The corresponding author attests that all listed authors meet authorship criteria and that no others meeting the criteria have been omitted. LM is the guarantor.

## TRANSPARENCY DECLARATION

The correspondent author affirms that the manuscript is an honest, accurate, and transparent account of the study being reported; that no important aspects of the study have been omitted; and that any discrepancies from the study as planned have been explained.

## DISSEMINATION TO PARTICIPANTS AND RELATED PATIENT AND PUBLIC COMMUNITIES

Results of this study have been made available to the public through an open access preprint posted to MedRxiv (doi: 10.1101/2020.11.27.20237966). The Spanish Society of Internal Medicine (SEMI) shares the results of the studies derived from the SEMI-COVID-19 Registry through its public facing website (https://www.fesemi.org/investigacion/proyectos/registro-semi-covid-19) and its twitter account (@Sociedad_SEMI).

## DATA SHARING

The data that support the findings of this study are available on request from the SEMI-COVID-19 Scientific Committee and the Registry Coordinating Center.

