## Supplemental Material for "Predicting critical illness on initial diagnosis of COVID-19 based on easily-obtained clinical variables: Development and validation of the PRIORITY model"

**Supplementary Table S1**. TRIPOD Checklist for Prediction Model Development and Validation.

**Supplementary Figure S1**. Selection of predictors using the least absolute shrinkage and selection operator (LASSO) method. (a) Tuning parameter (λ) selection from the LASSO model using 10-fold cross-validation via minimum criteria. The mean square error curve was plotted vs. log (λ). Dotted vertical lines at the optimal value by the minimum criteria, and the 1 standard error of the minimum criteria. Solid vertical line at the consensus criteria on the maximum number of predictors in the model. (b) LASSO coefficient profiles of the 21 potential predictors, plotted against the log (λ) sequence.

**Supplementary Figure S2.** Web-based calculator for predicting critical illness among patients with COVID-19. Accessible at https://www.evidencio.com/models/show/2344.

**Supplementary Figure S3**. Calibration curves of the model predicting COVID-19 critical illness. (a). AUC ROC in the development cohort, n=7850 patients from hospitals with equal or more than 300 beds. (b). AUC ROC in the validation cohort, n=2583 patients from hospitals with less than 300 beds. Upper, X-axis representing model predictions and y-axis observed critical illness rates. Circles representing deciles of risk according to the model predictions, plotted against the observed critical illness rate. Linear and local (LOESS) regression were represented to visualize the agreement between observed and predicted values. Below, histogram of predicted critical illness across the range of risk predictions.

**Supplementary Appendix S1**. List of the SEMI-COVID-19 network members.

This supplementary material has been provided by the authors to give readers additional information about their work.

**Supplementary Table S1. TRIPOD Checklist for Prediction Model Development and Validation.**

| **Section/Topic** | **Item** |  | **Checklist Item** | **Page** |
| --- | --- | --- | --- | --- |
| **Title and abstract** |  |  |  | |
| Title | 1 | D;V | Identify the study as developing and/or validating a multivariable prediction model, the target population, and the outcome to be predicted. | 1 |
| Abstract | 2 | D;V | Provide a summary of objectives, study design, setting, participants, sample size, predictors, outcome, statistical analysis, results, and conclusions. | 2-3 |
|  |  |  | Introduction | |
| Background and objectives | 3a | D;V | Explain the medical context (including whether diagnostic or prognostic) and rationale for developing or validating the multivariable prediction model, including references to existing models. | 4 |
|  | 3b | D;V | Specify the objectives, including whether the study describes the development or validation of the model or both. | 5 |
| **Methods** |  |  |  | |
| Source of data | 4a | D;V | Describe the study design or source of data (e.g., randomized trial, cohort, or registry data), separately for the development and validation data sets, if applicable. | 5 |
|  | 4b | D;V | Specify the key study dates, including start of accrual; end of accrual; and, if applicable, end of follow-up. | 5 |
| Participants | 5a | D;V | Specify key elements of the study setting (e.g., primary care, secondary care, general population) including number and location of centers. | 5 |
|  | 5b | D;V | Describe eligibility criteria for participants. | 5 |
|  | 5c | D;V | Give details of treatments received, if relevant. | N/A |
| Outcome | 6a | D;V | Clearly define the outcome that is predicted by the prediction model, including how and when assessed. | 6 |
|  | 6b | D;V | Report any actions to blind assessment of the outcome to be predicted. | N/A |
| Predictors | 7a | D;V | Clearly define all predictors used in developing or validating the multivariable prediction model, including how and when they were measured. | 6-7 |
|  | 7b | D;V | Report any actions to blind assessment of predictors for the outcome and other predictors. | N/A |
| Sample size | 8 | D;V | Explain how the study size was arrived at. | 5 |
| Missing data | 9 | D;V | Describe how missing data were handled (e.g., complete-case analysis, single imputation, multiple imputation) with details of any imputation method. | 8 |
| Statistical analysis methods | 10a | D | Describe how predictors were handled in the analyses. | 8-9 |
|  | 10b | D | Specify type of model, all model-building procedures (including any predictor selection), and method for internal validation. | 8-9 |
|  | 10c | V | For validation, describe how the predictions were calculated. | 9 |
|  | 10d | D;V | Specify all measures used to assess model performance and, if relevant, to compare multiple models. | 9 |
|  | 10e | V | Describe any model updating (e.g., recalibration) arising from the validation, if done. | N/A |
| Risk groups | 11 | D;V | Provide details on how risk groups were created, if done. | N/A |
| Development vs. validation | 12 | V | For validation, identify any differences from the development data in setting, eligibility criteria, outcome, and predictors. | 9 |
| **Results** |  |  |  | |
| Participants | 13a | D;V | Describe the flow of participants through the study, including the number of participants with and without the outcome and, if applicable, a summary of the follow-up time. A diagram may be helpful. | 10 |
|  | 13b | D;V | Describe the characteristics of the participants (basic demographics, clinical features, available predictors), including the number of participants with missing data for predictors and outcome. | Table 1 |
|  | 13c | V | For validation, show a comparison with the development data of the distribution of important variables (demographics, predictors and outcome). | 12 |
| Model development | 14a | D | Specify the number of participants and outcome events in each analysis. | 10,12 |
|  | 14b | D | If done, report the unadjusted association between each candidate predictor and outcome. | N/A |
| Model specification | 15a | D | Present the full prediction model to allow predictions for individuals (i.e., all regression coefficients, and model intercept or baseline survival at a given time point). | 11, Table 2 |
|  | 15b | D | Explain how to the use the prediction model. | 13, Table 3 |
| Model performance | 16 | D;V | Report performance measures (with CIs) for the prediction model. | 13 |
| Model-updating | 17 | V | If done, report the results from any model updating (i.e., model specification, model performance). | N/A |
| **Discussion** |  |  |  | |
| Limitations | 18 | D;V | Discuss any limitations of the study (such as nonrepresentative sample, few events per predictor, missing data). | 14-15 |
| Interpretation | 19a | V | For validation, discuss the results with reference to performance in the development data, and any other validation data. | 15 |
|  | 19b | D;V | Give an overall interpretation of the results, considering objectives, limitations, results from similar studies, and other relevant evidence. | 14-15 |
| Implications | 20 | D;V | Discuss the potential clinical use of the model and implications for future research. | 16 |
| **Other information** | |  |  | |
| Supplementary information | 21 | D;V | Provide information about the availability of supplementary resources, such as study protocol, Web calculator, and data sets. | 11, Appendix Figure 2. |
| Funding | 22 | D;V | Give the source of funding and the role of the funders for the present study. | 17, Acknowledgement |

**Supplementary Figure S1. Selection of predictors using the least absolute shrinkage and selection operator (LASSO) method.** (a) Tuning parameter (λ) selection from the LASSO model using 10-fold cross-validation via minimum criteria. The mean square error curve was plotted vs. log (λ). Dotted vertical lines at the optimal value by the minimum criteria, and the 1 standard error of the minimum criteria. Solid vertical line at the consensus criteria on the maximum number of predictors in the model. (b) LASSO coefficient profiles of the 21 potential predictors, plotted against the log (λ) sequence.

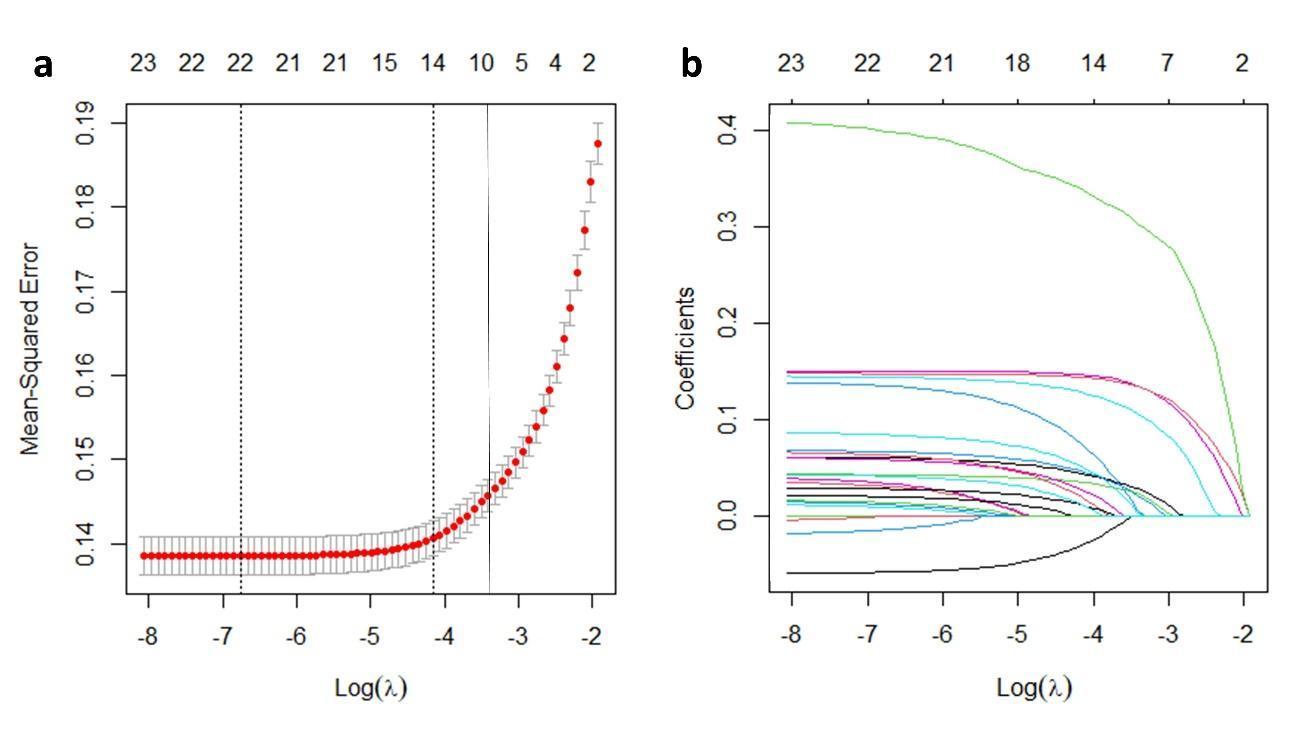

**Supplementary Figure S2. Web-based calculator for predicting critical illness among patients with COVID-19**. Accessible at https://www.evidencio.com/models/show/2344.

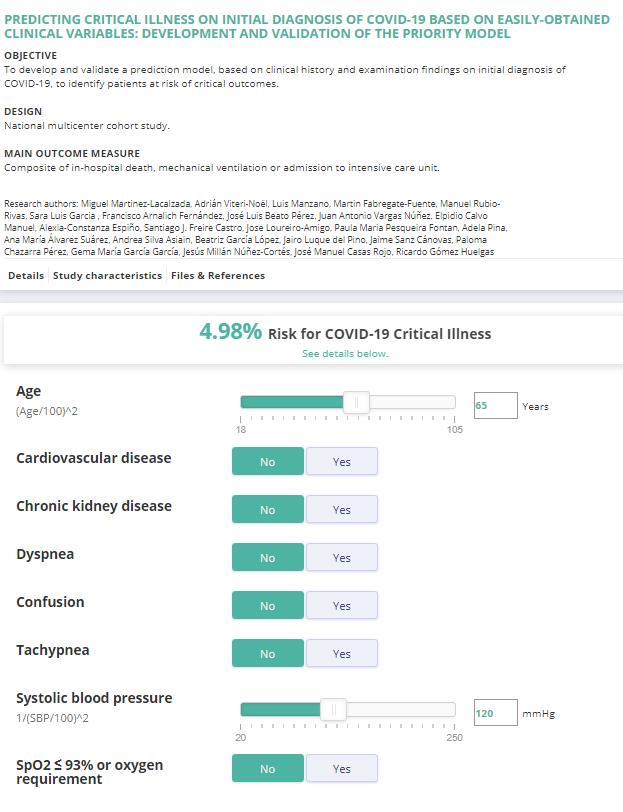

**Supplementary Figure S3. Calibration curves of the model predicting COVID-19 critical illness.** (a). AUC ROC in the development cohort, n=7850 patients from hospitals with equal or more than 300 beds. (b). AUC ROC in the validation cohort, n=2583 patients from hospitals with less than 300 beds. Upper, X-axis representing model predictions and y-axis observed critical illness rates. Circles representing deciles of risk according to the model predictions, plotted against the observed critical illness rate. Linear and local (LOESS) regression were represented to visualize the agreement between observed and predicted values. Below, histogram of predicted critical illness across the range of risk predictions.

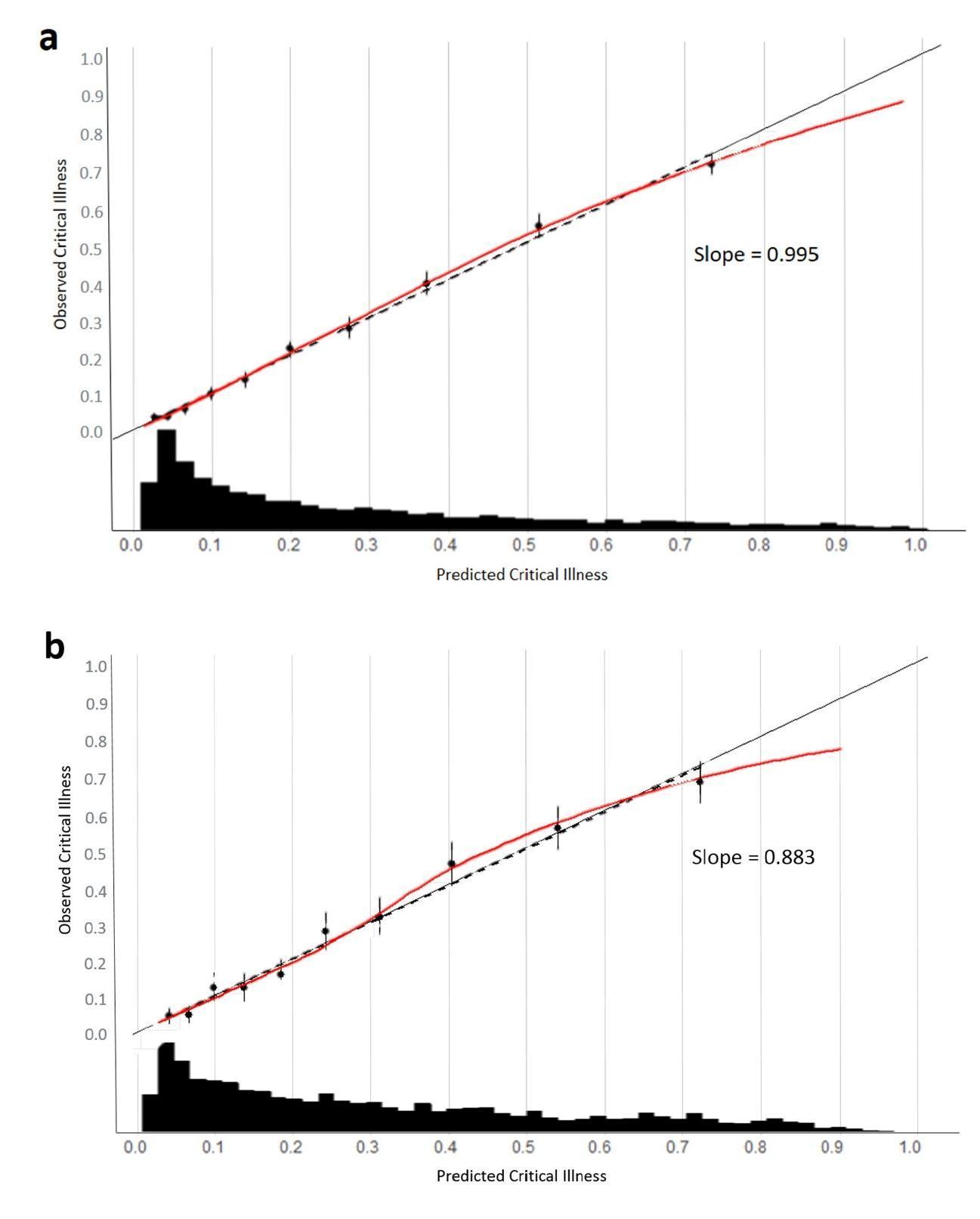

**Supplementary Appendix S1. LIST OF THE SEMI-COVID-19 NETWORK MEMBERS**

Coordinator of the SEMI-COVID-19 Registry: José Manuel Casas Rojo.

**SEMI-COVID-19 Scientific Committee Members**: José Manuel Casas Rojo, José Manuel Ramos Rincón, Carlos Lumbreras Bermejo, Jesús Millán Núñez-Cortés, Juan Miguel Antón Santos, Ricardo Gómez Huelgas.

**SEMI-COVID-19 Registry Coordinating Center**: S & H Medical Science Service.

**Members of the SEMI-COVID-19 Group:**

**Hospital Universitari de Bellvitge. L'Hospitalet de Llobregat.** Xavier Corbella, Narcís Homs, Abelardo Montero, Jose María Mora-Luján, Manuel Rubio-Rivas. **H. U. Gregorio Marañón. Madrid**. Laura Abarca Casas, Álvaro Alejandre de Oña, Rubén Alonso Beato, Leyre Alonso Gonzalo, Jaime Alonso Muñoz, Crhistian Mario Amodeo Oblitas, Cristina Ausín García, Marta Bacete Cebrián, Jesús Baltasar Corral, Maria Barrientos Guerrero, Alejandro Bendala Estrada, María Calderón Moreno, Paula Carrascosa Fernández, Raquel Carrillo, Sabela Castañeda Pérez, Eva Cervilla Muñoz, Agustín Diego Chacón Moreno, Maria Carmen Cuenca Carvajal, Sergio de Santos, Andrés Enríquez Gómez, Eduardo Fernández Carracedo, María Mercedes Ferreiro-Mazón Jenaro, Francisco Galeano Valle, Alejandra Garcia, Irene Garcia Fernandez-Bravo, María Eugenia García Leoni, Maria Gomez Antunez, Candela González San Narciso, Anthony Alexander Gurjian, Lorena Jiménez Ibáñez, Cristina Lavilla Olleros, Cristina Llamazares Mendo, Sara Luis García, Víctor Mato Jimeno, Clara Millán Nohales, Jesús Millán Núñez-Cortés, Sergio Moragón Ledesma, Antonio Muiño Miguez, Cecilia Muñoz Delgado, Lucía Ordieres Ortega, Susana Pardo Sánchez, Alejandro Parra Virto, María Teresa Pérez Sanz, Blanca Pinilla Llorente, Sandra Piqueras Ruiz, Guillermo Soria Fernández-Llamazares, María Toledano Macías, Neera Toledo Samaniego, Ana Torres do Rego, Maria Victoria Villalba Garcia, Gracia Villarreal, María Zurita Etayo. **H. U. La Paz-Cantoblanco-Carlos III. Madrid.** Jorge Álvarez Troncoso, Francisco Arnalich Fernández, Francisco Blanco Quintana, Carmen Busca Arenzana, Sergio Carrasco Molina, Aranzazu Castellano Candalija, Germán Daroca Bengoa, Alejandro de Gea Grela, Alicia de Lorenzo Hernández, Alejandro Díez Vidal, Carmen Fernández Capitán, Maria Francisca García Iglesias, Borja González Muñoz, Carmen Rosario Herrero Gil, Juan María Herrero Martínez, Víctor Hontañón, Maria Jesús Jaras Hernández, Carlos Lahoz, Cristina Marcelo Calvo, Juan Carlos Martín Gutiérrez, Monica Martinez Prieto, Elena Martínez Robles, Araceli Menéndez Saldaña, Alberto Moreno Fernández, Jose Maria Mostaza Prieto, Ana Noblejas Mozo, Carlos Manuel Oñoro López, Esmeralda Palmier Peláez, Marina Palomar Pampyn, Maria Angustias Quesada Simón, Juan Carlos Ramos Ramos, Luis Ramos Ruperto, Aquilino Sánchez Purificación, Teresa Sancho Bueso, Raquel Sorriguieta Torre, Clara Itziar Soto Abanedes, Yeray Untoria Tabares, Marta Varas Mayoral, Julia Vásquez Manau. **C. H. U. de Albacete. Albacete.** Jose Luis Beato Pérez, Maria Lourdes Sáez Méndez. **H. Clínico San Carlos. Madrid.** Inés Armenteros Yeguas, Javier Azaña Gómez, Julia Barrado Cuchillo, Irene Burruezo López, Noemí Cabello Clotet, Alberto E. Calvo Elías, Elpidio Calvo Manuel, Verónica Cano, Carmen María Cano de Luque, Cynthia Chocron Benbunan, Laura Dans Vilan, Claudia Dorta Hernández, Ester Emilia Dubon Peralta, Vicente Estrada Pérez, Santiago Fernandez-Castelao, Marcos Oliver Fragiel Saavedra, José Luis García Klepzig, Maria del Rosario Iguarán Bermúdez, Esther Jaén Ferrer, Alejandro Maceín Rodríguez, Alejandro Marcelles de Pedro, Rubén Ángel Martín Sánchez, Manuel Méndez Bailón, Sara Miguel Álvarez, Maria José Nuñez Orantos, Carolina Olmos Mata, Eva Orviz García, David Oteo Mata, Cristina Outon González, Juncal Perez-Somarriba, Pablo Pérez Mateos, Maria Esther Ramos Muñoz, Xabier Rivas Regaira, Laura Mª Rodríguez Gallardo, Iñigo Sagastagoitia Fornie, Alejandro Salinas Botrán, Miguel Suárez Robles, Maddalena Elena Urbano, Andrea María Vellisca González, Miguel Villar Martínez. **Hospital Royo Villanova. Zaragoza.** Nicolás Alcalá Rivera, Anxela Crestelo Vieitez, Esther del Corral Beamonte, Jesús Díez Manglano, Isabel Fiteni Mera, Maria del Mar Garcia Andreu, Martin Gerico Aseguinolaza, Claudia Josa Laorden, Raul Martínez Murgui, Marta Teresa Matía Sanz. **H. U. Puerta de Hierro. Majadahonda.** María Álvarez Bello, Ane Andrés Eisenhofer, Ana Arias Milla, Isolina Baños Pérez, Laura Benítez Gutiérrez, Javier Bilbao Garay, Silvia Blanco Alonso, Jorge Calderón Parra, Alejandro Callejas Díaz, José María Camino Salvador, Mª Cruz Carreño Hernández, Valentín Cuervas-Mons Martínez, Sara de la Fuente Moral, Miguel del Pino Jimenez, Alberto Díaz de Santiago, Itziar Diego Yagüe, Ignacio Donate Velasco, Ana María Duca, Pedro Durán del Campo, Gabriela Escudero López, Esther Expósito Palomo, Ana Fernández Cruz, Esther Fiz Benito, Andrea Fraile López, Amy Galán Gómez, Sonia García Prieto, Claudia García Rodríguez-Maimón, Miguel Ángel García Viejo, Javier Gómez Irusta, Edith Vanessa Gutiérrez Abreu, Isabel Gutiérrez Martín, Ángela Gutiérrez Rojas, Andrea Gutiérrez Villanueva, Jesús Herráiz Jiménez, Pedro Laguna del Estal, Mª Carmen Máinez Sáiz, Cristina Martín Martín, María Martínez Urbistondo, Fernando Martínez Vera, Susana Mellor Pita, Patricia Mills Sánchez, Esther Montero Hernández, Alberto Mora Vargas, Cristina Moreno López, Alfonso Ángel-Moreno Maroto, Victor Moreno-Torres Concha, Ignacio Morrás De La Torre, Elena Múñez Rubio, Ana Muñoz Gómez, Rosa Muñoz de Benito, Alejandro Muñoz Serrano, Jose María Palau Fayós, Lina Marcela Parra Ramírez, Ilduara Pintos Pascual, Arturo José Ramos Martín-Vegue, Antonio Ramos Martínez, Isabel Redondo Cánovas del Castillo, Alberto Roldán Montaud, Lucía Romero Imaz, Yolanda Romero Pizarro, Mónica Sánchez Santiuste, David Sánchez Órtiz, Enrique Sánchez Chica, Patricia Serrano de la Fuente, Pablo Tutor de Ureta, Ángela Valencia Alijo, Mercedes Valentín-Pastrana Aguilar, Juan Antonio Vargas Núñez, Jose Manuel Vázquez Comendador, Gema Vázquez Contreras, Carmen Vizoso Gálvez. **H. Miguel Servet. Zaragoza.** Gonzalo Acebes Repiso, Uxua Asín Samper, María Aranzazu Caudevilla Martínez, José Miguel García Bruñén, Rosa García Fenoll, Jesús Javier González Igual, Laura Letona Giménez, Mónica Llorente Barrio. **H. U. La Princesa. Madrid.** María Aguilera García, Ester Alonso Monge, Jesús Álvarez Rodríguez, Claudia Alvarez Varela, Miquel Berniz Gòdia, Marta Briega Molina, Marta Bustamante Vega, Jose Curbelo, Alicia de las Heras Moreno, Ignacio Descalzo Godoy, Alexia Constanza Espiño Alvarez, Ignacio Fernández Martín-Caro, Alejandra Franquet López-Mosteiro, Gonzalo Galvez Marquez, María J. García Blanco, Yaiza García del Álamo Hernández, Clara García-Rayo Encina, Noemí Gilabert González, Carolina Guillamo Rodríguez, Nicolás Labrador San Martín, Manuel Molina Báez, Carmen Muñoz Delgado, Pedro Parra Caballero, Javier Pérez Serrano, Laura Rabes Rodríguez, Pablo Rodríguez Cortés, Carlos Rodriguez Franco, Emilia Roy-Vallejo, Monica Rueda Vega, Aresio Sancha Lloret, Beatriz Sánchez Moreno, Marta Sanz Alba, Jorge Serrano Ballester, Alba Somovilla, Carmen Suarez Fernández, Macarena Vargas Tirado, Almudena Villa Marti. **H. U. de A Coruña. A Coruña.** Alicia Alonso Álvarez, Olaya Alonso Juarros, Ariadna Arévalo López, Carmen Casariego Castiñeira, Ana Cerezales Calviño, Marta Contreras Sánchez, Ramón Fernández Varela, Santiago J. Freire Castro, Ana Padín Trigo, Rafael Prieto Jarel, Fátima Raad Varea, Ignacio Ramil Freán, Laura Ramos Alonso, Francisco Javier Sanmartín Pensado, David Vieito Porto. **Hospital Clínico de Santiago. Santiago de Compostela.** Maria del Carmen Beceiro Abad, Maria Aurora Freire Romero, Sonia Molinos Castro, Emilio Manuel Paez Guillan, María Pazo Nuñez, Paula Maria Pesqueira Fontan. **H. de Cabueñes. Gijón.** Ana María Álvarez Suárez, Carlos Delgado Vergés, Rosa Fernandez-Madera Martínez, Eva Fonseca Aizpuru, Alejandro Gómez Carrasco, Cristina Helguera Amezua, Juan Francisco López Caleya, María del Mar Martínez López, Aleida Martínez Zapico, Carmen Olabuenaga Iscar, María Luisa Taboada Martínez, Lara María Tamargo Chamorro. **H. Moisès Broggi. Sant Joan Despí.** Judit Aranda Lobo, Jose Loureiro Amigo, Isabel Oriol Bermúdez, Melani Pestaña Fernández, Nicolas Rhyman, Nuria Vázquez Piqueras. **Hospital Universitario Dr. Peset. Valencia**. Juan Alberto Aguilera Ayllón, Arturo Artero, María del Mar Carmona Martín, María José Fabiá Valls, Maria de Mar Fernández Garcés, Ana Belén Gómez Belda, Ian López Cruz, Manuel Madrazo López, Elisabeth Mateo Sanchis, Jaume Micó Gandia, Laura Piles Roger, Adela Maria Pina Belmonte, Alba Viana García. **H. U. Ramón y Cajal. Madrid.** Luis Fernando	Abrego Vaca, Ana Andréu Arnanz, Octavio Arce García, Marta Bajo González, Pablo Borque Sanz, Alberto Cozar Llisto, Sonia de Pedro Baena, Beatriz Del Hoyo Cuenda, María Alejandra Gamboa Osorio, Isabel García Sánchez, Andrés González García, Oscar Alberto López Cisneros, Luis Manzano, Miguel Martínez-Lacalzada, Borja Merino Ortiz, Jimena Rey-García, Elisa Riera González, Cristina Sánchez Díaz, Grisell Starita Fajardo, Cecilia Suárez Carantoña, Adrian Viteri Noel, Svetlana Zhilina Zhilina. **H. Nuestra Señora del Prado. Talavera de la Reina**. Sonia Casallo Blanco, Jeffrey Oskar Magallanes Gamboa, Cristina Salazar Mosteiro, Andrea Silva Asiain. **C. Asistencial de Zamora. Zamora.** Carlos Aldasoro Frias, Luis Arribas Perez, María Esther Fraile Villarejo, Beatriz García López, Victor Madrid Romero, Emilia Martínez Velado, Victoria Palomar Calvo, Sara Pintos Otero, Carlota Tuñón de Almeida. **H. Virgen de la Salud. Toledo.** Ana Maria Alguacil Muñoz, Marta Blanco Fernández, Veronica Cano, Ricardo Crespo Moreno, Fernando Cuadra Garcia-Tenorio, Blanca Díaz-Tendero Nájera, Raquel Estévez González, María Paz García Butenegro, Alberto Gato Díez, Verónica Gómez Caverzaschi, Piedad María Gómez Pedraza, Julio González Moraleja, Raúl Hidalgo Carvajal, Patricia Jiménez Aranda, Raquel Labra González, Áxel Legua Caparachini, Pilar Lopez Castañeyra, Agustín Lozano Ancin, Jose Domingo Martin Garcia, Cristina Morata Romero, María Jesús Moya Saiz, Helena Moza Moríñigo, Gemma Muñiz Nicolás, Enriqueta Muñoz Platon, Filomena Oliveri, Elena Ortiz Ortiz, Raúl Perea Rafael, Pilar Redondo Galán, María Antonia Sepulveda Berrocal, Vicente Serrano Romero de Ávila, Pilar Toledano Sierra, Yamilex Urbano Aranda, Jesús Vázquez Clemente, Carmen Yera Bergua. **Hospital Costa del Sol. Marbella.** Javier García Alegría, Nicolás Jiménez-García, Jairo Luque del Pino, María Dolores Martín Escalante. **H. U. Infanta Cristina. Parla.** Juan Miguel Antón Santos, Ana Belén Barbero Barrera, Coralia Bueno Muiño, Ruth Calderón Hernaiz, Irene Casado Lopez, José Manuel Casas Rojo, Andrés Cortés Troncoso, Mayte de Guzmán García-Monge, Francesco Deodati, Gonzalo García Casasola Sánchez, Elena Garcia Guijarro, Davide Luordo, María Mateos González, Jose A Melero Bermejo, Lorea Roteta García, Elena Sierra Gonzalo, Javier Villanueva Martínez. **Hospital Regional Universitario de Málaga. Málaga.** Mª Mar Ayala Gutiérrez, Rosa Bernal López, José Bueno Fonseca, Verónica Andrea Buonaiuto, Luis Francisco Caballero Martínez, Lidia Cobos Palacios, Clara Costo Muriel, Francis de Windt, Ana Teresa Fernandez-Truchaud Christophel, Paula García Ocaña, Ricardo Gómez Huelgas, Javier Gorospe García, Maria Dolores López Carmona, Pablo López Quirantes, Almudena López Sampalo, Elizabeth Lorenzo Hernández, Juan José Mancebo Sevilla, Jesica Martin Carmona, Luis Miguel Pérez-Belmonte, Araceli Pineda Cantero, Carlos Romero Gómez, Michele Ricci, Jaime Sanz Cánovas. **H. U. San Juan de Alicante. San Juan de Alicante**. Marisa Asensio Tomás, David Balaz, David Bonet Tur, Ruth Cañizares Navarro, Paloma Chazarra Pérez, Jesús Corbacho Redondo, Eliana Damonte White, Leticia Espinosa Del Barrio, Pedro Jesús Esteve Atiénzar, Carles García Cervera, David Francisco García Núñez, Vicente Giner Galvañ, Angie Gómez Uranga, Javier Guzmán Martínez, Isidro Hernández Isasi, Lourdes Lajara Villar, Verónica Martínez Sempere, Juan Manuel Núñez Cruz, Sergio Palacios Fernández, Juan Jorge Peris García, Andrea Riaño Pérez, José Miguel Seguí Ripoll, Azucena Sempere Mira, Philip Wikman-Jorgensen. **H. del Henares. Coslada.** Jesús Ballano Rodríguez-Solís, Luis Cabeza Osorio, María del Pilar Fidalgo Montero, Mª Isabel Fuentes Soriano, Erika Esperanza Lozano Rincon, Ana Martín Hermida, Jesus Martinez Carrilero, Jose Angel Pestaña Santiago, Manuel Sánchez Robledo, Patricia Sanz Rojas, Nahum Jacobo Torres Yebes, Vanessa Vento. **H. U. La Fe. Valencia.** Dafne Cabañero, María Calabuig Ballester, Pascual Císcar Fernández, Ricardo Gil Sánchez, Marta Jiménez Escrig, Cristina Marín Amela, Laura Parra Gómez, Carlos Puig Navarro, José Antonio Todolí Parra. **H. de Mataró. Mataró.** Raquel Aranega González, Ramon Boixeda, Javier Fernández Fernández, Carlos Lopera Mármol, Marta Parra Navarro, Ainhoa Rex Guzmán, Aleix Serrallonga Fustier. **H. San Pedro. Logroño.** Diana Alegre González, Irene Ariño Pérez de Zabalza, Sergio Arnedo Hernández, Jorge Collado Sáenz, Beatriz Dendariena, Marta Gómez del Mazo, Iratxe Martínez de Narvajas Urra, Sara Martínez Hernández, Estela Menendez Fernández, Jose Luís Peña Somovilla, Elisa Rabadán Pejenaute. **Complejo Hospitalario Universitario Ourense. Ourense.** Raquel Fernández González, Amara Gonzalez Noya, Carlos Hernández Ceron, Isabel Izuzquiza Avanzini, Ana Latorre Diez, Pablo López Mato, Ana María Lorenzo Vizcaya, Daniel Peña Benítez, Milagros María Peña Zemsch, Lucía Pérez Expósito, Marta Pose Bar, Lara Rey González, Laura Rodrigo Lara. **H. Juan Ramón Jiménez. Huelva.** Francisco Javier Bejarano Luque, Francisco Javier Carrasco-Sánchez, Mercedes de Sousa Baena, Jaime Díaz Leal, Aurora Espinar Rubio, Maria Franco Huertas, Juan Antonio García Bravo, Andrés Gonzalez Macías, Encarnación Gutiérrez Jiménez, Alicia Hidalgo Jiménez, Constantino Lozano Quintero, Carmen Mancilla Reguera, Francisco Javier Martínez Marcos, Francisco Muñoz Beamud, Maria Perez Aguilera, Alícia Perez Jiménez, Virginia Rodríguez Castaño, Alvaro Sánchez de Alcazar del Río, Leire Toscano Ruiz. **H. U. Reina Sofía. Córdoba.** Antonio Pablo Arenas de Larriva, Pilar Calero Espinal, Javier Delgado Lista, Francisco Fuentes-Jiménez, María Jesús Gómez Vázquez, Jose Jiménez Torres, José López-Miranda, Laura Martín Piedra, Javier Pascual Vinagre, Pablo Pérez-Martinez, María Elena Revelles Vílchez, Juan Luis Romero Cabrera, José David Torres Peña. **Hospital Infanta Margarita. Cabra.** María Esther Guisado Espartero, Lorena Montero Rivas, Maria de la Sierra Navas Alcántara, Raimundo Tirado-Miranda. **Hospital Alto Guadalquivir.** Andújar. Begoña Cortés Rodríguez. **C. H. U. de Ferrol. Ferrol.** Hortensia Alvarez Diaz, Tamara Dalama Lopez, Estefania Martul Pego, Carmen Mella Pérez, Ana Pazos Ferro, Sabela Sánchez Trigo, Dolores Suarez Sambade, Maria Trigas Ferrin, Maria del Carmen Vázquez Friol, Laura Vilariño Maneiro**. C. H. U. de Badajoz. Badajoz.** Rafael Aragon Lara, Inmaculada Cimadevilla Fernandez, Juan Carlos Cira García, Gema Maria García García, Julia Gonzalez Granados, Beatriz Guerrero Sánchez, Francisco Javier Monreal Periáñez, Maria Josefa Pascual Perez. **Complejo Asistencial Universitario de León. León.** Rosario Maria García Diez, Manuel Martin Regidor, Angel Luis Martínez Gonzalez, Alberto Muela Molinero, Raquel Rodríguez Díez, BeatrizVicente Montes. **Hospital Marina Baixa. Villajoyosa.** Javier Ena, Jose Enrique Gómez Segado. **Hospital del Tajo. Aranjuez** Ruth Gonzalez Ferrer, Raquel Monsalvo Arroyo. **H. de Pozoblanco. Pozoblanco.** José Nicolás Alcalá Pedrajas, Antonia Márquez García, Inés Vargas. **H. U. Marqués de Valdecilla. Santander.** Marta Fernández-Ayala Novo, José Javier Napal Lecumberri, Nuria Puente Ruiz, Jose Riancho, Isabel Sampedro Garcia. **H. U. Severo Ochoa. Leganés.** Yolanda Casillas Viera, Lucía Cayuela Rodríguez, Carmen de Juan Alvarez, Gema Flox Benitez, Laura García Escudero, Juan Martin Torres, Patricia Moreira Escriche, Susana Plaza Canteli, M Carmen Romero Pérez. **Hospital Platón. Barcelona**. Ana Suarez Lombraña. **Hospital Valle del Nalón. Riaño. Langreo**. Sara Fuente Cosío, César Manuel Gallo Álvaro, Julia Lobo García, Antía Pérez Piñeiro. **H. General Defensa.** Anyuli Gracia Gutiérrez, Leticia Esther Royo Trallero. **H. U. del Vinalopó. Elche.** Francisco Amorós Martínez, Erika Ascuña Vásquez, José Carlos Escribano Stablé, Adriana Hernández Belmonte, Ana Maestre Peiró, Raquel Martínez Goñi, M.Carmen Pacheco Castellanos, Bernardino Soldan Belda, David Vicente Navarro. **H. Francesc de Borja. Gandia.** Alba Camarena Molina, Simona Cioaia, Anna Ferrer Santolalia, José María Frutos Pérez, Eva Gil Tomás, Leyre Jorquer Vidal, Marina Llopis Sanchis, M Ángeles Martínez Pascual, Alvaro Navarro Batet, Mari Amparo Perea Ribis, Ricardo Peris Sanchez, José Manuel Querol Ribelles, Silvia Rodriguez Mercadal, Ana Ventura Esteve. **H. G. U. de Castellón. Castellón de la Plana.** Jorge Andrés Soler, Marián Bennasar Remolar, Alejandro Cardenal Álvarez, Daniela Díaz Carlotti, María José Esteve Gimeno, Sergio Fabra Juana, Paula García López, María Teresa Guinot Soler, Daniela Palomo de la Sota, Guillem Pascual Castellanos, Ignacio Pérez Catalán, Celia Roig Martí, Paula Rubert Monzó, Javier Ruiz Padilla, Nuria Tornador Gaya, Jorge Usó Blasco. **C. A. U. de Salamanca. Salamanca.** Gloria María Alonso Claudio, Víctor Barreales Rodríguez, Cristina Carbonell Muñoz, Adela Carpio Pérez, María Victoria Coral Orbes, Daniel Encinas Sánchez, Sandra Inés Revuelta, Miguel Marcos Martín, José Ignacio Martín González, José Ángel Martín Oterino, Leticia Moralejo Alonso, Sonia Peña Balbuena, María Luisa Pérez García, Ana Ramon Prados, Beatriz Rodríguez-Alonso, Ángela Romero Alegría, Maria Sanchez Ledesma, Rosa Juana Tejera Pérez. **Hospital Doctor José Molina Orosa. Arrecife Lanzarote. Virginia Herrero García, Berta Román Bernal. H. U. del Sureste. Arganda del Rey**. Jon Cabrejas Ugartondo, Ana Belén Mancebo Plaza, Arturo Noguerado Asensio, Bethania Pérez Alves, Natalia Vicente López. **Hospital de Palamós. Palamós.** Ana Alberich Conesa, Mari Cruz Almendros Rivas, Miquel Hortos Alsina, José Marchena Romero, Anabel Martin-Urda Diez-Canseco. **H. U. Quironsalud Madrid. Pozuelo de Alarcón. Madrid**. Pablo Guisado Vasco, Ana Roda Santacruz, Ana Valverde Muñoz. **H. Parc Tauli. Sabadell.** Francisco Epelde, Isabel Torrente. **H. Virgen de los Lirios. Alcoy. Alicante**. Mª José Esteban Giner. **Hospital Clínico Universitario de Valladolid. Valladolid.** Xjoylin Teresita Egües Torres, Sara Gutiérrez González, Cristina Novoa Fernández, Pablo Tellería Gómez. **H. G. U. de Elda. Elda.** Carmen Cortés Saavedra, Jennifer Fernández Gómez, Borja González López, María Soledad Hernández Garrido Ana Isabel López Amorós, Santiago López Gil, Maria de los Reyes Pascual Pérez, Nuria Ramírez Perea, Andrea Torregrosa García. **Hospital Público de Monforte de Lemos. Monforte de Lemos.** José López Castro, Manuel Lorenzo López Reboiro. **H. Virgen del Mar. Madrid.** Thamar Capel Astrua, Paola Tatiana Garcia Giraldo, Maria Jesús González Juárez, Victoria Marquez Fernandez, Ada Viviana Romero Echevarry**. Hospital do Salnes. Vilagarcía de Arousa.** Vanesa	Alende Castro, Ana María Baz Lomba, Ruth Brea Aparicio, Marta Fernandez Morales, Jesús Manuel Fernández Villar, María Teresa López Monteagudo, Cristina Pérez García, Lorena Rodríguez Ferreira, Diana Sande Llovo, Maria Begoña Valle Feijoo. **Hospital Quironsalud A Coruña. A Coruña.** Hector Meijide Miguez. **H. U. 12 de Octubre. Madrid.** Paloma Agudo de Blas, Coral Arévalo Cañas, Blanca Ayuso, José Bascuñana Morejón, Samara Campos Escudero, María Carnevali Frías, Santiago Cossio Tejido, Borja de Miguel Campo, Carmen Díaz Pedroche, Raquel Diaz Simon, Ana García Reyne, Lucia Jorge Huerta, Antonio Lalueza Blanco, Jaime Laureiro Gonzalo, Jaime Lora-Tamayo, Carlos Lumbreras Bermejo, Guillermo Maestro de la Calle, Barbara Otero Perpiña, Diana Paredes Ruiz, Marcos Sánchez Fernández, Javier Tejada Montes.
